# Neonatal Paenibacilliosis: *Paenibacillus thiaminolyticus* as a Novel Cause of Neonatal Sepsis with High Risk of Sequelae in Uganda

**DOI:** 10.1101/2022.11.07.22281940

**Authors:** Jessica E. Ericson, Kathy Burgoine, Elias Kumbakumba, Moses Ochora, Christine Hehnly, Francis Bajunirwe, Joel Bazira, Claudio Fronterre, Cornelia Hagmann, Abhaya V. Kulkarni, M. Senthil Kumar, Joshua Magombe, Edith Mbabazi-Kabachelor, Sarah U. Morton, Mercedeh Movassagh, John Mugamba, Ronald Mulondo, Davis Natukwatsa, Brian Nsubuga Kaaya, Peter Olupot-Olupot, Justin Onen, Kathryn Sheldon, Jasmine Smith, Paddy Ssentongo, Peter Ssenyonga, Benjamin Warf, Emmanuel Wegoye, Lijun Zhang, Julius Kiwanuka, Joseph N. Paulson, James R. Broach, Steven J. Schiff

## Abstract

*Paenibacillus thiaminolyticus* may be an underdiagnosed cause of neonatal sepsis. We prospectively enrolled a cohort of 800 neonates presenting with a clinical diagnosis of sepsis at two Ugandan hospitals. Quantitative polymerase chain reaction specific to *P. thiaminolyticus* and to the *Paenibacillus* genus were performed on the blood and cerebrospinal fluid (CSF) of 631 neonates who had both specimen types available. Neonates with virus detected in either specimen type were considered to potentially have paenibacilliosis, (37/631, 6%). We described antenatal, perinatal, and neonatal characteristics, presenting signs, and 12-month developmental outcomes for neonates with paenibacillosis vs. clinical sepsis. Median age at presentation was 3 (interquartile range 1, 7) days. Fever (92%), irritability (84%) and seizures (51%) were common. Eleven (30%) had an adverse outcome: 5 (14%) neonates died during the first year of life; 5 of 32 (16%) survivors developed postinfectious hydrocephalus and one (3%) additional survivor had neurodevelopmental impairment without hydrocephalus. These results highlight the need to consider local pathogen prevalence and the possibility of unusual pathogens when determining antibiotic choice for neonatal sepsis.

---

Neonatal sepsis is a leading cause of early childhood death worldwide.^1,2^ Affected infants are disproportionately from low-resource settings.^2^ Survivors of neonatal sepsis have an increased risk of neurodevelopmental impairment^3,4^, hydrocephalus^5^ and cerebral palsy.^6^ Effective antibiotic therapy relies on identification of the causative pathogen or, in the absence of this information, empirical therapy that is broad enough to provide effective coverage of the most likely pathogens.^6^

In low-resource settings, blood and cerebrospinal fluid cultures are often unavailable or uninformative.^7^ Cultures can be negative for a variety of reasons including low sample volume and technical limitations.^8^ Some pathogens are unculturable or are difficult to culture using routine culture methods. Culture-independent methods to identify causative pathogens have recently become possible due to advances in molecular diagnostics. Because of their cost and technical requirements, these methods are not widely available and have only rarely been used to identify pathogens affecting neonates in low-resource settings.^9,10^

Current international guidelines recommend the combination of ampicillin and gentamicin as first-line empirical antibiotic therapy for neonates with sepsis.^11^ However, in regions where antibiotic resistance is common, these antibiotics are not the ideal treatment for many neonatal infections.^12-14^ Without local culture and antibiotic susceptibility testing, the risk of antibiotic resistance is unknown; in these cases, ampicillin and gentamicin may be inadequate treatment. ^15,16 15,16 15,16 15,16 15,16^

In previous work, using targeted metagenomics, we found 41% of infants with postinfectious hydrocephalus, a common sequela of NS in Uganda, had a *Paenibacillus spp* infection.^15,17^ We sought to describe the clinical syndrome of neonatal *Paenibacillus* infection among Ugandan neonates presenting with clinical signs of sepsis, to report 12-month outcomes for infants with this novel infection and to compare patient characteristic and outcomes for infants with *Paenibacillus* infection compared to infants with clinical sepsis without *Paenibacillus* detected. We hypothesized that infants with *Paenibacillus* detected would be more likely to have the composite outcome of postinfectious hydrocephalus, death or neurodevelopmental impairment than those without *Paenibacillus* detected.

## Methods

### Study Population

Ugandan neonates (≤ 28 days of age) previously enrolled in a parent study were evaluated for inclusion in this subanalysis focused on Paenibacillus infection. The parent study enrolled 800 neonates presenting to two regional referral hospitals with clinical signs of neonatal sepsis (fever, poor feeding and lethargy; hypothermia, poor feeding, and lethargy; seizures and/or bulging fontanelle, poor feeding, and fever) were recruited. The study sites were, Mbale Regional Referral Hospital in Eastern Uganda, and Mbarara Regional Referral Hospital in Western Uganda. Neonates born at a gestational age < 37 weeks or < 2000 grams birthweight, and those who had been diagnosed with birth asphyxia or hypoxic ischemic encephalopathy were excluded. Following informed written consent, blood and cerebrospinal fluid were collected from each neonate using aseptic technique. Infants were eligible for this subanalysis if they had sufficient blood and CSF to perform qPCR for *Paenibacillus* genus and *P. thiaminolyticus* (N=631).

### Laboratory Analysis

An aliquot of each was collected into DNA/RNA preservative (DNA/RNA Shield, Zymo Corporation) and frozen at -80° C. Additional aliquots of blood and CSF were processed in the local clinical laboratory for standard-of-care clinical tests. Once the entire cohort had been enrolled, the frozen samples were transferred to Penn State University for processing. CSF was available for 631/800 (79%) of the neonates enrolled. For these 631, we performed quantitative polymerase chain reaction (qPCR) on the blood and CSF using primers specific to both the *Paenibacillus* genus and *P. thiaminolyticus*. Neonates with detection of virus using either test were considered to have possible paenibacilliosis.

Demographics, birth history, clinical signs at presentation and during the hospital stay, results of laboratory tests performed as part of clinical care and antibiotics administered were abstracted from each infant’s medical record. In Uganda, current clinical guidelines recommend that the umbilical cord stump receive no care (dry cord care) or be cleansed with chlorhexidine (Umbigel™). Mothers were asked to indicate how they cared for the neonate’s cord stump at home including any substances applied to the cord stump prior to presentation. The presence of seizure-like movements was considered to represent seizures; no electroencephalograms were performed. Fever was defined as a temperature ≥38° C.

Infants underwent developmental assessments at 2 months, 6 months and 12 months of age using the *Bayley Scales of Infant and Toddler Development Third Edition* (BSID-III).^18^ Population-normalized BSID-III scores range from 1 to 19 with a population mean of 10, and a standard deviation (SD) of 3. Neurodevelopmental impairment (NDI) was defined as a BSID-III score < -2SD on any of the subscales.

We compared the demographic characteristics, presenting clinical signs, antibiotic treatment and clinical course for patients with neonatal paenibacillosis to septic neonates without *P. thiaminolyticus* detected using Fisher’s exact test for proportions and Wilcoxon rank-sum test for continuous variables. Similarly, we compared the proportion of patients with and without paenibacilliosis who experienced in-patient death, infant death (death between birth and 1 year of age), postinfectious hydrocephalus (PIH), NDI, and the composite of infant death, PIH or NDI during the first 12 months of age using Fisher’s exact test. Finally, for infants with paenibacilliosis, we compared the proportion of patients with fever, irritability, seizure, tachypnea, respiratory distress, umbilical cord discharge, hypertonia, tachycardia, bulging fontanelle and stiff neck between the two study sites. Missingness was uncommon and no adjustment or imputation was undertaken.

Neonates with PIH were referred to CURE Children’s Hospital of Uganda, a neurosurgical specialty hospital located in Mbale for further management. Additional specimens of CSF were collected as part of the neurosurgical evaluation. Each specimen was split into two aliquots: one that was collected into DNA/RNA preservative (DNA/RNA Shield, Zymo Corporation) and frozen at -80° C and one that was frozen without preservative. An additional 61 infants with PIH had their CSF processed in the same manner. The fresh frozen samples were thawed and 1 mL of CSF from each patient was inoculated into a BD BACTEC lytic anaerobic medium blood culture bottle supplemented with 1 mL of defibrinated horse blood (Thermo Scientific). Culture bottles were incubated in a BD BACTECTM FX instrument and monitored for bacterial growth for up to 14 days. Culture bottles that were positive for bacterial growth were subcultured on BD BBLTM Chocolate II and CDC Anaerobe 5% Sheep Blood agar plates and incubated at 37° C under anaerobic conditions (Anoxomat, Advanced Instruments). Culture bottles that remained negative after 14 days were also subcultured under anaerobic conditions though none of these resulted in bacterial growth. All subsequent culturing after initial anaerobic conditions was done aerobically. Three CSF samples were positive for growth in culture bottles. As previously described, colonies from subculture plates were used for Gram stain, organism identification by MALDI-TOF, biochemical testing, and antimicrobial susceptibility testing. Biochemical testing was performed using API 50 CH strip following manufacturers protocol. Susceptibility testing and interpretations were performed by E-test method using Clinical and Laboratory Standards Institute (CLSI) guidelines.

This study was approved by the Human Subjects Protection Program at The Pennsylvania State University, Pennsylvania, United States, by CURE Children’s Hospital of Uganda Institutional Review Board, the Mbarara University of Science and Technology Research Ethics Committee, and with oversight of the Ugandan National Council on Science and Technology. Informed written consent was obtained from each patient’s mother prior to enrollment. All data produced in the current study are available upon reasonable request to the authors and will be made publicly available once the parent neonatal sepsis study is published.

## Results

### Clinical Epidemiology of Paenibacillosis

Six percent (37/631) of neonates with clinical sepsis had PCR evidence of *Paenibacillus* infection. CSF PCR was positive in most cases, 35/631 (6%) patients; 1/35 (3%) of these also had a positive blood PCR. Two additional patients had a positive blood PCR but negative CSF PCR (Supplemental Table 1).

**Table 1.** Demographics of neonates with clinical sepsis due with and without *Paenibacillus thiaminolyticus* detected by qPCR.

|  | Paenibacillus<br>PCR Positive<br>N=37 (6%) | Paenibacillus<br>PCR Negative<br>N=594 (94%) | P |
| --- | --- | --- | --- |
| Median age at sepsis presentation, days (25<sup>th</sup>,<br>75<sup>th</sup> percentiles) | 3 (1, 7) | 2 (1, 4) | 0.129 |
| <3 days | 22 (61) | 433 (73) |  |
| 3-7 days | 6 (17) | 55 (9) |  |
| 8-14 days | 7 (19) | 46 (8) |  |
| >14 days | 1 (3) | 59 (10) |  |
| Median gestational age at birth, weeks (25<sup>th</sup>,<br>75<sup>th</sup> percentiles) | 39 (38, 40) | 40 (39, 41) | 0.266 |
| Median birth weight, g (25<sup>th</sup>, 75<sup>th</sup> percentiles) | 3200 (3000,<br>3500) | 3200 (2900,<br>3500) | 0.505 |
| Sex |  |  |  |
| Female | 20 (54) | 335 (56) | 0.865 |
| Male | 17 (46) | 259 (44) |  |
| Maternal HIV status |  |  | 0.209 |
| Positive | 4 (11) | 29 (5) |  |
| Negative | 31 (84) | 539 (91) |  |
| Unknown | 2 (5) | 27 (5) |  |
| Median maternal age, years (25<sup>th</sup>, 75<sup>th</sup><br>percentiles) | 26 (23, 30) | 24 (21, 29) | 0.443 |
| Median maternal parity (25<sup>th</sup>, 75<sup>th</sup> percentiles) | 2 (1, 4) | 2 (1, 4) | 0.402 |
| Maternal fever during pregnancy | 19 (51) | 333 (51) | 0.608 |
| Maternal fever during labor | 8 (22) | 164 (28) | 0.487 |
| Delivery location |  |  |  |
| Healthcare | 29 (78) | 555 (94) | 0.003* |
| Hospital | 21 (57) | 422 (71) |  |
| Health center | 7 (19) | 113 (19) |  |
| Clinic | 1 (3) | 20 (3) |  |
| Home | 8 (22) | 36 (6) |  |
| Delivery mode |  |  | 0.166 |
| Vaginal | 27 (73) | 361 (61) |  |
| C-section | 10 (27) | 230 (39) |  |
| Rupture of membranes >18 hours | 8 (24) | 101 (19) | 0.495 |
| Umbilical cord care |  |  |  |
| None | 23 (62) | 475 (81) | 0.011† |
| Any substance | 14 (38) | 113 (19) |  |
| Saliva | 6 (16) | 41 (7) |  |
| Cosmetic | 6 (16) | 35 (6) |  |
| Plant material | 2 (5) | 28 (5) |  |
| Other | 0 (0) | 9 (10) |  |
| Feeding method |  |  | 0.123 |
| Exclusively breast fed | 34 (92) | 561 (94) |  |
| Breast milk and water | 1 (3) | 1 (<1) |  |
| Breast milk and replacement feeding | 1 (3) | 20 (3) |  |
| Replacement feeding | 1 (3) | 4 (1) |  |
| Unknown | 0 (0) | 8 (1) |  |
| Location |  |  | 0.085 |
| Mbarara | 21 (57) | 244 (41) |  |
| Mbale | 16 (43) | 350 (59) |  |

\*Fisher's exact test comparing the proportion of patients with and without *Paenibacillus* detected who were born at home vs. in a healthcare facility (clinic, health centre or hospital); †Fisher's exact test comparing the proportion of patients with and without *Paenibacillus* detected who had any vs. no substances applied to the umbilical cord stump.

Postnatal age at presentation was similar for neonates with and without paenibacillosis, median age 3 (IQR 1, 7) days and 2 (1, 4) days, respectively, P=0.129 (Table 1). Most neonates with paenibacillosis were born vaginally (73%) in a healthcare facility (hospital, lower level health center or private clinic) (78%). However, when compared to infants without paenibacilliosis, infants with paenibacilliosis detected were significantly more likely to be born at home, 6% vs. 22%, respectively, P<0.01 and to have received non-recommended cord stump care, 19% vs. 38%, respectively, P=0.01.

Neonates with paenibacilliosis frequently had fever (92%) and irritability (84%) (Figure 1). Clinical seizures were present in half of neonates. A bulging fontanelle was present in 22% overall and in 6/16 (38%) infants at Mbale but in only 2/21 (10%) at Mbarara, P=0.06. All other presenting signs occurred in a similar proportion of neonates at each of the two sites.

**Figure 1.**
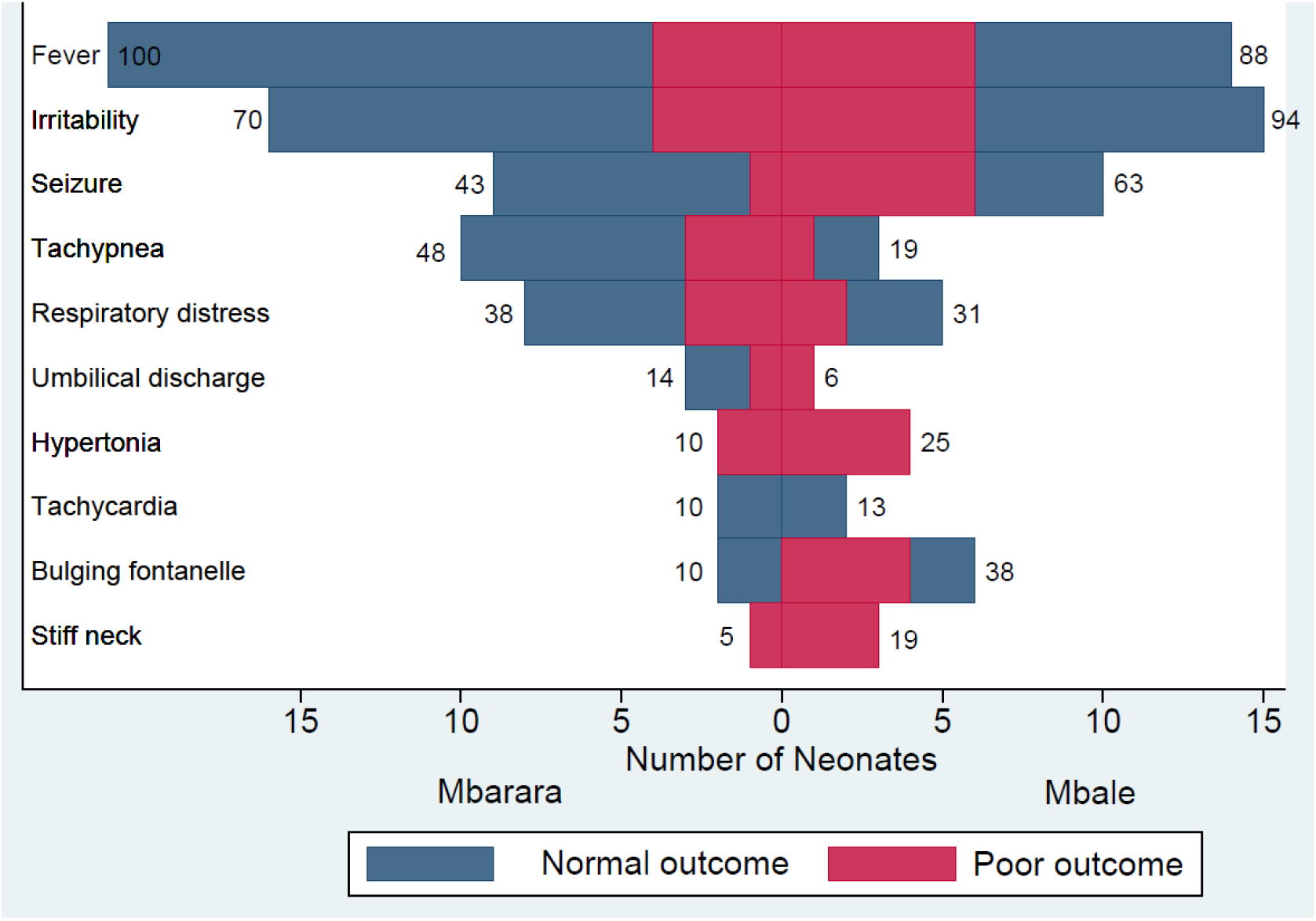
Presenting signs and outcomes for neonates with *Paenibacillus thiaminolyticus* detected by qPCR during clinical sepsis. Number of neonates presenting to each site with each sign of infection. Numbers at the end of each bar indicate the proportion of neonates at each site who had the corresponding sign of infection. Neonates with the composite poor outcome of death, postinfectious hydrocephalus or moderate/severe neurodevelopmental impairment are indicated in red.

### Laboratory studies

Blood cultures grew an organism in 4/37 (14%) of neonates (Supplemental Table 2). Two grew *Staphylococcus aureus*, and one each grew a *Klebsiella species*, a *Bacillus species* and *Streptococcus agalactiae*. Due to testing limitations at the local microbiology laboratory, species-level identification of some organisms was not performed. One of the neonates with *S. aureus* died shortly after hospital admission. All 37 neonates had negative antigen testing for malaria. Thirty-six patients had a blood CMV PCR performed; none was positive.

**Table 2.** Description of neonates who developed postinfectious hydrocephalus following neonatal sepsis with *P. thiaminolyticus* detected by qPCR

|  | 1 | 2 | 3 | 4 | 5 |
| --- | --- | --- | --- | --- | --- |
| Age at presentation | 8-14 days | < 3 days | 8-14 days | 3-7 days | 3-7 days |
| Sex | Female | Male | Male | Male | Male |
| Birth location | Home | Home | Home | Home | Home |
| Cord care | Cosmetic powder | Cosmetic powder | Vaseline | None | None |
| Presenting signs | Fever, poor feeding, irritability, bulging fontanelle, lethargy, seizure, umbilical discharge | Fever, poor feeding, irritability, bulging fontanelle, lethargy, seizure, hypertonia | Fever, poor feeding, irritability, bulging fontanelle, lethargy, respiratory distress, hypotonia | Fever, poor feeding, irritability, lethargy, vomiting, seizure, hypotonia | Fever, poor feeding, irritability, lethargy, seizure, stiff neck, hypertonia |
| CSF cell count, 10<sup>6</sup> cells/L | ≤5 | 75 | 80 | ≤5 | ≤5 |
| CSF protein, g/L | 3.4 | 1.8 | 0.5 | 1 | 2.3 |
| Blood culture result | Negative | Negative | Negative | Negative | Negative |
| Admission heart rate, beats/minute | 125 | 148 | 138 | 136 | 131 |
| Admission respiratory rate, breaths/minute | 36 | 51 | 67 | 59 | 47 |
| Initial antibiotic treatment | 3 days<br>ampicillin/gentamicin, | 2 days ampicillin/gentamicin | 2 days<br>ampicillin/gentamicin | 7 days<br>ceftriaxone/gentamicin | 9 days<br>ampicillin/gentamicin |
| Additional antibiotic treatment | 11 days<br>ceftriaxone/gentamicin | 7 days ceftriaxone/gentamicin, 14 days ceftriaxone/amikacin<br>14 days enteral amoxicillin/ciprofloxacin/metronidazole | 4 days<br>cefotaxime/gentamicin, 14 days ceftriaxone/amikacin |  |  |
| Hydrocephalus treatment | None | Shunted | Shunted | None | None |
| 6-month outcome | Moderate cognitive and motor impairment | Not seen | Moderate cognitive and motor impairment | Normal | Not seen |
| 12-month outcome | Not seen | Moderate cognitive and motor impairment | Not seen | Normal | Not seen |

Seventeen of the 37 paenibacilliosis patients had a CSF WBC count reported. Four of the 17 (24%) neonates with an available CSF WBC count had a CSF WBC count ≥ 15: 20, 75, 75 and 80 ×10^6^ cells/L. The protein concentration was ≥ 1 g/L in two of these four (50%) neonates (1.8, 5.4 g/L) and in an additional 6 neonates (8/37, 22%) with normal or unavailable CSF WBC counts (1, 1, 1, 2.3, 3.4, 5 g/L). CSF and blood glucose concentrations were not available. No CSF culture grew bacteria in the local laboratory.

### Treatment

The choice of empirical therapy was chosen by the admitting healthcare worker and therefore varied across the cohort. Most neonates with paenibacilliosis (26/37, 70%) were initially started on intravenous ampicillin plus gentamicin. Eight (21%) were treated initially with a third-generation cephalosporin (ceftriaxone or cefotaxime) along with either gentamicin (5, 13%), or an aminoglycoside, ampicillin and cloxacillin (2, 7%). Three (8%) received ampicillin, cloxacillin and gentamicin as initial therapy and 1 (3%) received ampicillin, gentamicin and ceftriaxone.

Antibiotic therapy was escalated to a broader antibiotic regimen for 9/37 (24%) neonates. Antibiotic escalation occurred after a median of 3 days (IQR: 3, 3). This is common practice in these hospitals, when the treating clinician does not see a good clinical response. Most commonly, ampicillin was changed to a cephalosporin (5/9, 56%). In two cases each (2/9, 22%), cloxacillin was added to ampicillin and gentamicin and in 1 case (1/9, 11’%) gentamicin was changed to a cephalosporin and ampicillin was continued.

### Outcomes

The composite poor outcome of infant death, PIH or NDI was more common in neonates with paenibacilliosis than those without, 11/37 (30%) vs. 79/594 (13%), P=0.012. Among neonates with paenibacilliosis, there was no difference in the frequency of the composite poor outcome between the two sites, 7/16 (44%) at Mbale and 4/21 (19%) at Mbarara, P=0.151 (Figure 2). Infant death following paenibacilliosis occurred in 5/37 (14%) and occurred in a similar proportion of neonates cared for at each site, 2/16 (13%) at Mbale and 3/21 (14%) at Mbarara, P=1.00. Three patients progressed rapidly to death prior to hospital discharge. Another infant remained critically ill during the hospital stay and was discharged to home against medical advice; this infant died shortly after discharge home. A fifth patient was treated with 5 days of ampicillin and gentamicin, was discharged home from the hospital in good condition but developed a second febrile illness and died at 2 months of age.

**Figure 2.**
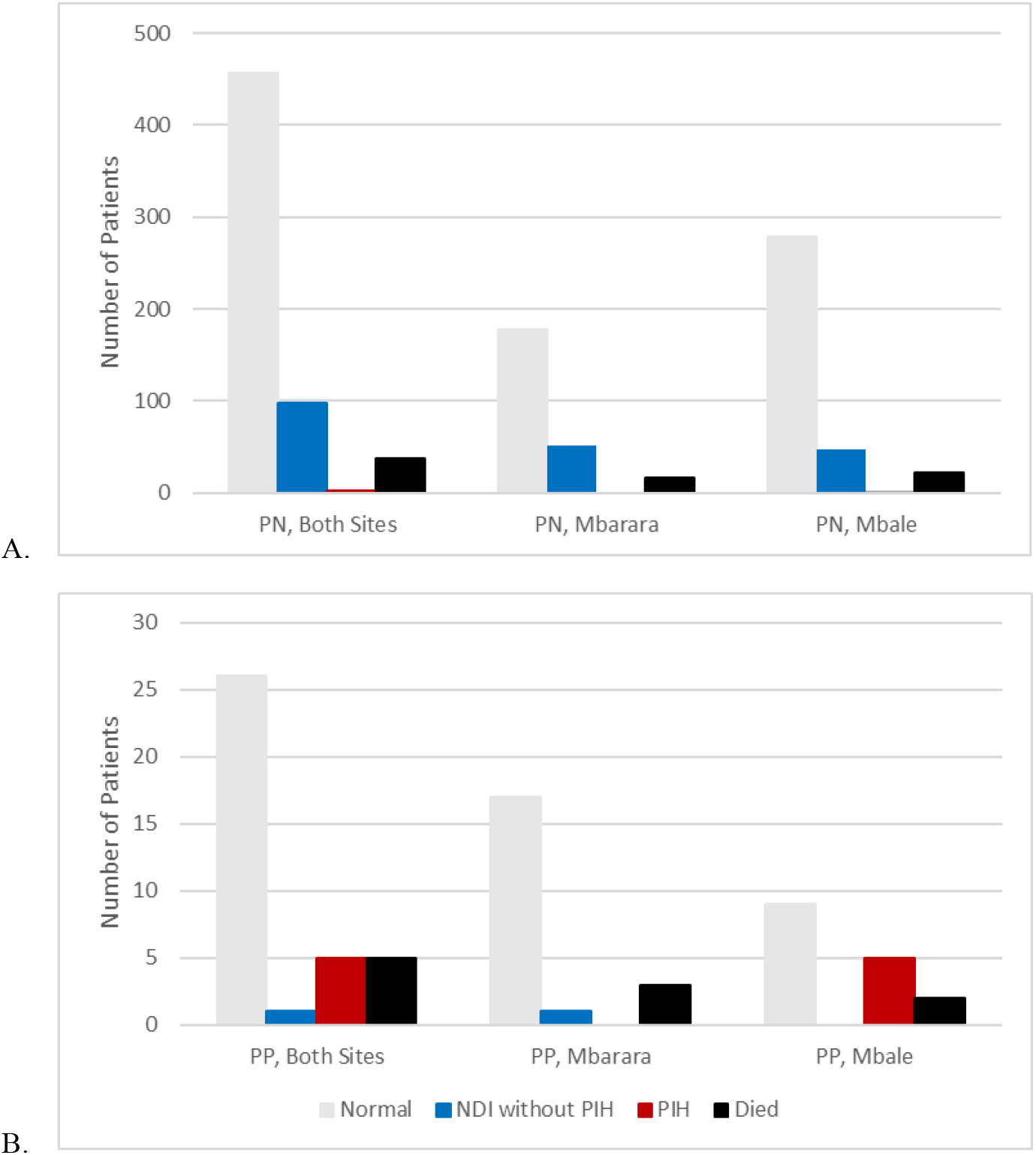
Outcomes for neonates with clinical sepsis with *Paenibacillus* negative (PN) and *Paenibacillus* positive (PP) qPCR results for *Paenibacillus thiaminolyticus* detection in the CSF at both sites and at Mbarara and Mbale.

PIH was also more common among neonates with paenibacilliosis than those neonates without paenibacilliosis, 5/37 (14%) vs. 3/594 (<1%), P<0.001. Additionally, PIH occurred following paenibacilliosis more frequently at Mbale, 5/16 (31%), than at Mbarara, 0/21 (0%), P=0.010. Four of the five (80%) neonates with *Paenibacillus*-associated PIH had an elevated CSF protein concentration but all 5/5 (100%) had a CSF WBC count < 100 ×10^6^ cells/L at presentation (Table 2). Two required placement of a ventricular peritoneal shunt, two were managed conservatively without surgery and one was referred for neurosurgical evaluation but was lost to follow-up. Nineteen of the 32 (59%) paenibacilliosis survivors had a developmental assessment performed at 6 months and 22/32 (69%) at 12 months; 29 (91%) had a developmental assessment at 6 or 12 months of age. Three of the 4 patients with paenibacilliosis-associated PIH who remained in care had moderate/severe neurodevelopmental impairment as assessed at 6- or 12-months of age (Table 3). One additional neonate (4%) who survived without hydrocephalus had NDI at the last developmental assessment.

**Table 3.** Developmental outcomes for survivors of neonatal sepsis with *P. thiaminolyticus* detected by qPCR

|  | **6 months**<br>**N=19 (%)** | **12 months**<br>**N=22 (%)** | **Last follow up**<br>**N=29 (%)** |
| --- | --- | --- | --- |
| Scaled cognitive score, mean ± SD | 11 ± 5 | 13 ± 3 | 13 ± 5 |
| Moderate/severe impairment | 2 (11) | 1 (5) | 3 (10) |
| Scaled fine motor score, mean ± SD | 12 ± 5 | 12 ± 3 | 12 ± 5 |
| Moderate/severe impairment | 2 (11) | 1 (5) | 2 (7) |
| Scaled gross motor score, mean ± SD | 11 ± 4 | 10 ± 4 | 9 ± 4 |
| Moderate/severe impairment | 2 (11) | 2 (9) | 4 (14) |
| Scaled receptive communication score, mean ± SD | 10 ± 4 | 10 ± 3 | 10 ± 4 |
| Moderate/severe impairment | 2 (11) | 1 (5) | 2 (7) |
| Scaled expressive communication score, mean ± SD | 9 ± 4 | 10 ± 2 | 10 ± 3 |
| Moderate/severe impairment | 2 (11) | 0 (0) | 3 (10) |

The three isolates of *P. thiaminolyticus* that were successfully cultured from infants with PIH following NS were all non-susceptible to vancomycin (Table 4). Two of the three (67%) were resistant to ampicillin. There are not interpretive criteria for the clinical significance of minimum inhibitory concentrations (MIC) for ceftriaxone but 2 of the 3 had a very low MIC and the third had an MIC of 0.25 ug/mL, a concentration which is probably susceptible.^19^

**Table 4.** E-test Antibiotic susceptibility testing results for *P. thiaminolyticus* isolates obtained from 3 infants with post-infectious hydrocephalus as a sequela of neonatal sepsis.

|  | Mbale |  | Mbale2 |  | Mbale3 |  |
| --- | --- | --- | --- | --- | --- | --- |
|  | MIC (µg/mL) | Interpretation | MIC (µg/mL) | Interpretation | MIC (µg/mL) | Interpretation |
| Ampicillin | 0.12 | S | 1 | R | 0.5 | R |
| Ceftriaxone | 0.03 | ** | 0.25 | ** | 0.06 | ** |
| Ciprofloxacin | 0.12 | S | 0.12 | S | 0.12 | S |
| Clindamycin | 1 | I | 1 | I | 4 | R |
| Gentamicin | 2 | S | 2 | S | 2 | S |
| Penicillin | 0.12 | S | 0.5 | R | 0.25 | R |
| Meropenem | 0.25 | S | 0.5 | S | 0.5 | S |
| Tetracycline | 0.5 | S | 2 | S | 2 | S |
| Vancomycin | 8 | Nonsusceptible | 8 | Nonsusceptible | 8 | Nonsusceptible |

MIC: minimum inhibitory concentration; S: susceptible; R: resistant; I: intermediate

## Discussion

We describe the first cohort of neonates with sepsis due to *Paenibacillus species*; most infections were due to *P. thiaminolyticus*. Signs of meningitis such as irritability, seizures and bulging fontanelle were common at presentation. Eleven percent of neonates died during their original in-patient admission. Poor outcomes were common among survivors: PIH developed in 16% and NDI was common.

*P. thiaminolyticus* has rarely been reported as a cause of human disease. The first case of human infection was reported in 2008 when an 80-year-old man undergoing hemodialysis developed bacteremia due to *P. thiaminolyticus*.^20^ He received 4 weeks of vancomycin and improved. A 33-year-old Swiss woman experienced a surgical wound infection due to *P. thiaminolyticus* 7 days following an abdominoplasty procedure.^21^ The organism was identified on culture of an aspirate from an abdominal wall fluid collection. She was treated with 2 weeks of unspecified intravenous antibiotics followed by 2 weeks of amoxicillin-clavulanate as definitive therapy and completely recovered. Finally, *P. thiaminolyticus* was recovered on blood culture from a 25-day old neonate admitted to a hospital in the United States of America due to cardiorespiratory arrest following 1 day of poor feeding and increased sleep.^22^ Unfortunately, the neonate succumbed to her infection 4 days later. Post-mortem examination revealed a soft brain with several areas of infarction but without clear signs of meningitis. Our finding that 33 of 631 (5%) neonates evaluated for sepsis had *P. thiaminolyticus* detected using molecular methods was unexpected and suggests that *P. thiaminolyticus* may be an underdiagnosed cause of neonatal sepsis, meningitis, and PIH in Uganda.

Studies seeking to identify causative organisms for neonatal sepsis may fail to identify *P. thiaminolyticus* for several reasons. First, this organism may have ecological niches that are not universally distributed. We failed to find any evidence of *P. thiaminolyticus* presence in the vaginal microbiome of 99 women residing in Mbale or Mbarara, Uganda at the time of delivery suggesting that neonates may become colonized with the organism through environmental, rather than maternal sources.^23^ It has been identified in fish from Lake Michigan in the United States,^24^ and from the soil in India.^25^ Other species of *Paenibacillus* have been identified in the soil globally^26-30^ and as a member of the human gut microbiome.^31^ Some species are known to infect honeybees^32^ and, rarely, humans.^33-35^ Neonates cared for in industrialized or high-resource settings may have limited contact with environmental reservoirs of *Paenibacillus*. Studies evaluating neonatal pathogens in high-resource settings may fail to identify *Paenibacillus* because there are no neonatal infections in those cohorts caused by *Paenibacillus*.

Neonates who are born and live in environments where exposure to contaminated water and soil is common could encounter *Paenibacillus* more frequently and at higher levels. It is possible that neonatal infections due to *Paenibacillus* do occur globally but are not diagnosed in either high- or low-resource settings due to the limitations of culture-based pathogen detection methods in neonates. Even common neonatal pathogens can be missed when the blood volume used to inoculate the blood culture bottles is <1 mL.^36,37^ Low sample volume will similarly have limited our ability to detect *Paenibacillus* in an infant with paenibacillosis. Blood cultures are typically incubated for 5-7 days and CSF cultures for 2-5 days. *Paenibacillus* may take 7-14 days to reach the level of detection using standard culture methods.^38^ Cultures may be deemed negative and the specimens discarded prior to sufficient time to detect *Paenibacillus* elapsing. Studies attempting to diagnose the spectrum of pathogens causing disease in neonatal sepsis may be improved by increasing the duration of specimen incubation beyond the standard 5-7 days or by using molecular methods to improve diagnostic yield.

In our prior work, we were able to grow *P. thiaminolyticus* from the CSF of 3 neonates with PIH by inoculating the CSF into anaerobic lytic blood culture bottles. Anaerobic blood culture bottles are not used as routine part of the evaluation of neonatal sepsis in most clinical settings but have been shown to increase the diagnostic yield of blood cultures for neonates with bacteremia.^39^ Culture sensitivity for fastidious organisms such as *Kingella kingae* is also known to be improved when synovial fluid is incubated in a blood culture bottle.^40,41^ The addition of an anaerobic blood culture bottle to the CSF evaluation of neonates with sepsis may improve the diagnosis of infections due to *Paenibacillus* and facultative anaerobes implicated in neonatal sepsis. Using these methods in all neonates with sepsis may not be warranted if the prevalence of infections identified using them is low. However, in research settings that attempt to describe the range of organisms infecting young infants expanding culture-based techniques to include additional types of media could potentially reduce false negatives and allow for a more complete description of the causative bacterial agents.

All of the neonates in our cohort with paenibacillosis were diagnosed using molecular methods. The addition of a *Paenibacillus* PCR to the evaluation of neonatal sepsis could improve the diagnosis of this emerging infection by providing a rapid, low-biomass way of identifying affected neonates. PCR and other molecular methods have been shown to be useful for the diagnosis of other fastidious organisms that cause disease in neonates such as *Mycoplasma hominis*,^42^ *Ureaplasma parvum*,^43^ *Leptotrichia amnionii*,^44^ and *Sneathia amnii*.^45^ Epidemiological studies of neonatal serious bacterial infections that only use culture-based methods likely underestimate both the number and variety of infections caused by bacteria.

The diagnosis of *Paenibacillus* infection is important for the care of neonates with sepsis due to the high incidence of associated mortality and morbidity. In contrast to infections due *Streptococcus agalactiae* (0%^46^-4%^47^), paenibacillosis resulted in PIH in 12% of cases. Meningitis due to *E. coli* causes PIH in 18^47^-22%^46^. In our cohort, *Paenibacillus* conferred a similar risk of PIH as *E. coli* but was a more common cause of infection in our cohort (data not shown). Thus, paenibacillosis may be a leading cause of PIH in Uganda. It is unknown whether optimal treatment of neonatal paenibacillosis would reduce the incidence of PIH or NDI in the region. We assessed NDI using the BSID-III. Although the BSID-III uses age-based normative values generated from an American population, the tool has been shown to be a valid means of comparing development for young African infants to each other but not to infants from other regions of the world.^48,49^ We found that NDI occurred following clinical neonatal sepsis in a similar proportion of infants with and without paenibacilliosis.

The World Health Organization recommends using the combination of ampicillin and gentamicin as the empirical antibiotic regimen for neonatal sepsis due to the coverage provided against *Streptococcus agalactiae* (Group B streptococcus), *Escherichia coli*, and *Listeria monocytogenes*.^50^ Regions where antibiotic resistance is common or additional organisms are typical may need to use a broader spectrum regimen empirically.^51,52^ Our discovery that *P. thiaminolyticus* could be a common cause of culture-negative sepsis in Uganda has important implications for empiric antibiotic selection. Antibiotic susceptibility testing performed on the three isolates we successfully isolated from neonates with PIH demonstrated resistance to ampicillin and vancomycin. We additionally identified the presence of several beta-lactamase genes which could confer resistance to ampicillin.^53^ Prior studies have found that resistance to ampicillin, vancomycin, clindamycin and tetracycline is common in *Paenibacillus* species.^21,54^ Because there are few clinical reports of *P. thiaminolyticus*, it is unknown how often resistance occurs or what antibiotic regimens would maximize the likelihood of a good outcome.

Inappropriate empirical antibiotic therapy has been associated with worse outcomes for neonatal sepsis. ^55-57^ These studies assume that an appropriate antibiotic is ultimately administered within 1-3 days of symptom onset.^58^ Unfortunately, *P. thiaminolyticus* can be difficult to recover using culture and will be consistently missed in Uganda and other parts of the world where neonatal sepsis is common.^59,60^ Thus, it is likely that many infected neonates do not receive effective antimicrobial therapy at all, thus increasing the risk of mortality and serious sequelae, including PIH. Incorporating the possibility of a difficult-to-culture organism into antibiotic guidelines and protocols has the potential to improve outcomes. Our findings suggest that the combination of a third-generation cephalosporin and gentamicin would be preferred over ampicillin and gentamicin as an empiric antibiotic regimen for neonatal sepsis in Uganda especially when meningitis is suspected. A prior study of soil *Paenibacillus species’* suggests that ∼70% of isolates are susceptible to ceftriaxone, an antibiotic that has good penetration into the central nervous system.^54^ Gentamicin is less reliable at achieving therapeutic concentrations in the central nervous system, especially if an abscess is present, but resistance appears to be quite uncommon.^54,61^ Antibiotic susceptibility testing should be performed to guide antibiotic management in individual cases of paenibacilliosis. Performing antibiotic susceptibility testing on a larger number of isolates will be important to improve our understanding of how best to care for neonates with these infections.

The possibility of antibiotic resistance should be considered when antibiotics are chosen to treat a neonate with paenibacilliosis.^62^ In particular, the WHO-recommended first-line antibiotic regimen for the treatment of neonatal sepsis (ampicillin plus gentamicin), is unlikely to be effective for *P. thiaminolyticus*. Vancomycin is commonly used for gram positive central nervous system (CNS) infections and is the drug-of-choice for CNS infections due to *Bacillus species* but should not be used empirically for treatment of *Paenibacillus* infections.^63^ It is possible that alternate antibiotic regimens may be optimal for treatment of this novel infection. Because this organism may be an important pathogen in the developing world, any antibiotic consideration would need to achieve therapeutic concentrations in the CNS, be inexpensive, easily administered, and well tolerated with few side effects.

Nutritional interventions may also be important adjunctive treatments for *P. thiaminolyticus* infections. This organism produces thiaminase, an uncommon bacterial enzyme which has the potential to reduce thiamine levels.^24^ Recent studies have failed to show a mortality benefit of the combination of thiamine, vitamin C with and without hydrocortisone in mortality outcomes for adults with sepsis.^64-66^ However, in a retrospective analysis, thiamine supplementation improved survival for adults with hospital-acquired pneumonia.^67^ Thiamine supplementation may yet have a role for certain types of infection, in cases when sepsis is caused by a thiamine-consuming organism or when pre-existing thiamine deficiency exists.^68-70^ Furthermore, thiamine supplementation may improve neurodevelopmental outcomes or reduce the likelihood of other sequelae, such as PIH, following neonatal sepsis even if it does not infer a survival benefit.^71,72^ Studies evaluating the role of thiamine supplementation as adjunctive therapy for neonatal sepsis due to thiaminase-producing *P. thiaminolyticus* are needed.

An important area of future research will be to define the qPCR level that is clinically significant. In this report, we included all neonates who had any *Paenibacillus* result. It is possible that some infants included had infections due to other organisms, or even non-infectious reasons for presenting with clinical signs consistent with infection. Additionally, the outcomes that we present following neonatal paenibacilliosis may reflect inadequate antibiotic therapy being more common among neonates with *Paenibacillus* detected than those without. Infants with paenibacilliosis may be more likely to receive ineffective antibiotic therapy than infants without paenibacilliosis. This could explain some of the increased frequency of poor outcomes in infants with *Paenibacillus* detected. Since the causative pathogen was unknown in the majority of non-paenibacilliosis cases, we were not able to assess antibiotic-microbe concordance in order to assess for this possibility.

## Conclusion

*Paenibacillus species* was identified in 6% of neonates with signs of sepsis who presented to two Ugandan referral hospitals; most of these were *P. thiaminolyticus*. Improved diagnostics for neonatal sepsis are urgently needed in the region. Optimal antibiotic treatment for this infection is unknown but ampicillin and vancomycin will be ineffective in many cases. The role of thiamine supplementation as adjunctive therapy is unknown but rational to consider.

## Supporting information

STROBE Checklist

## Data Availability

All data produced in the current study are available upon reasonable request to the authors and will be made publicly available once the parent neonatal sepsis study is published.

## Acknowledgements

This study and the parent neonatal sepsis study were funded by the National Institutes of Health under 1DP1HD086071(S.J.S) and 1R01AI145057 (S.J.S). J.E.E. was supported by National Center for Advancing Translational Sciences grant #KL2 TR002015.

---

**Supplemental Table 1.** *Paenibacillus* genus and *Paenibacillus thiaminolyticus* qPCR results from blood and cerebrospinal fluid.

|  | Cerebrospinal Fluid |  | Blood |  |
| --- | --- | --- | --- | --- |
| Subject | *Paenibacillus*<br>genus | *Paenibacillus*<br>*thiaminolyticus* | *Paenibacillus*<br>genus | *Paenibacillus*<br>*thiaminolyticus* |
| 1 | 51,846 | 909 | ND | ND |
| 2 | 419,144 | 926,763 | ND | ND |
| 3 | 540,471 | 905,393 | ND | ND |
| 4 | 335 | 509 | ND | ND |
| 5 | ND | 255,011 | ND | ND |
| 6 | 55 | 1749 | ND | ND |
| 7 | 62 | ND | ND | ND |
| 8 | 118 | ND | ND | ND |
| 9 | 147 | ND | ND | ND |
| 10 | 151 | ND | ND | ND |
| 11 | 152 | 1585 | ND | ND |
| 12 | 178 | 1217 | ND | ND |
| 13 | 204 | ND | ND | ND |
| 14 | 208 | 2048 | ND | ND |
| 15 | 212 | 2225 | ND | ND |
| 16 | 240 | ND | ND | ND |
| 17 | 245 | 1606 | ND | ND |
| 18 | 280 | ND | ND | ND |
| 19 | 324 | ND | ND | ND |
| 20 | 363 | 3704 | ND | ND |
| 21 | 462 | 4537 | ND | ND |
| 22 | 742 | 1930 | ND | ND |
| 23 | 955 | 1171 | ND | ND |
| 24 | 1077 | 2848 | ND | ND |
| 25 | 1094 | 6549 | ND | ND |
| 26 | 1456 | ND | ND | ND |
| 27 | 22,561 | 1427 | ND | ND |
| 28 | 27,897 | 1437 | ND | ND |
| 29 | 30,023 | 1011 | ND | ND |
| 30 | 32,109 | 43,556 | ND | ND |
| 31 | 68,263 | 2505 | ND | ND |
| 32 | 140,435 | 12,387 | 5846 | 12,541 |
| 33 | 173,836 | ND | ND | ND |
| 34 | 300,260 | ND | ND | ND |
| 35 | ND | 827 | ND | ND |
| 36 | ND | ND | ND | 699 |
| 37 | ND | ND | ND | 214 |

ND: not detected

**Supplemental Table 2.**
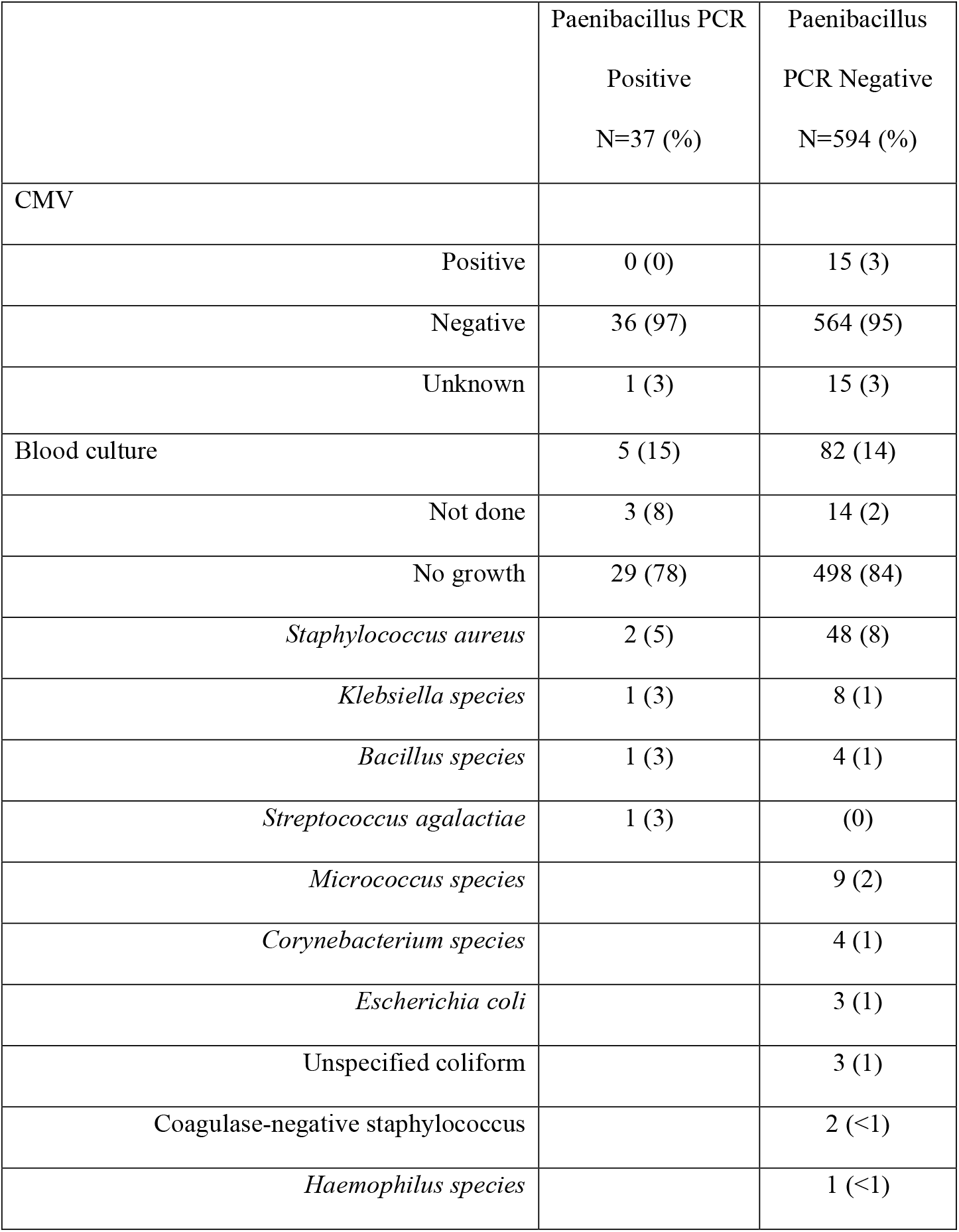
Results of blood polymerase chain reaction tests for cytomegalovirus and blood cultures for neonates with and without *Paenibacillus* detected.

|  | Paenibacillus PCR<br>Positive<br>N=37 (%) | Paenibacillus<br>PCR Negative<br>N=594 (%) |
| --- | --- | --- |
| CMV |  |  |
| Positive | 0 (0) | 15 (3) |
| Negative | 36 (97) | 564 (95) |
| Unknown | 1 (3) | 15 (3) |
| Blood culture | 5 (15) | 82 (14) |
| Not done | 3 (8) | 14 (2) |
| No growth | 29 (78) | 498 (84) |
| *Staphylococcus aureus* | 2 (5) | 48 (8) |
| *Klebsiella species* | 1 (3) | 8 (1) |
| *Bacillus species* | 1 (3) | 4 (1) |
| *Streptococcus agalactiae* | 1 (3) | (0) |
| *Micrococcus species* |  | 9 (2) |
| *Corynebacterium species* |  | 4 (1) |
| *Escherichia coli* |  | 3 (1) |
| Unspecified coliform |  | 3 (1) |
| Coagulase-negative staphylococcus |  | 2 (<1) |
| *Haemophilus species* |  | 1 (<1) |

## Notes

### Competing Interest Statement

The authors have declared no competing interest.

### Author Declarations

The Human Subjects Protection Program at The Pennsylvania State University gave ethical approval for this work. The CURE Childrens Hospital of Uganda Institutional Review Board gave ethical approval for this work. The Mbarara University of Science and Technology Research Ethics Committee gave ethical approval for this work.

## References

1. Taylor AW, Blau DM, Bassat Q, et al. Initial findings from a novel population-based child mortality surveillance approach: a descriptive study. Lancet Glob Health 2020;8(7):e909–e919. (In eng). DOI: 10.1016/S2214-109X(20)30205-9.

2. Fleischmann C, Reichert F, Cassini A, et al. Global incidence and mortality of neonatal sepsis: a systematic review and meta-analysis. Arch Dis Child 2021 (In eng). DOI: 10.1136/archdischild-2020-320217.

3. Shim SY, Cho SJ, Park EA. Neurodevelopmental Outcomes at 18-24 Months of Corrected Age in Very Low Birth Weight Infants with Late-onset Sepsis. J Korean Med Sci 2021;36(35):e205. (In eng). DOI: 10.3346/jkms.2021.36.e205.

4. Shadbolt R, We MLS, Kohan R, et al. Neonatal Staphylococcus Aureus Sepsis: a 20-year Western Australian experience. J Perinatol 2022 (In eng). DOI: 10.1038/s41372-022-01440-3.

5. Warf BC, Dagi AR, Kaaya BN, Schiff SJ. Five-year survival and outcome of treatment for postinfectious hydrocephalus in Ugandan infants. J Neurosurg Pediatr 2011;8(5):502–8. (In eng). DOI: 10.3171/2011.8.PEDS11221.

6. Al-Matary A, Al Sulaiman M, Al-Otaiby S, Qaraqei M, Al-Matary M. Association between the timing of antibiotics administration and outcome of neonatal sepsis. J Infect Public Health 2022;15(6):643–647. (In eng). DOI: 10.1016/j.jiph.2022.05.004.

7. Saha SK, Schrag SJ, El Arifeen S, et al. Causes and incidence of community-acquired serious infections among young children in south Asia (ANISA): an observational cohort study. Lancet 2018;392(10142):145–159. (In eng). DOI: 10.1016/S0140-6736(18)31127-9.

8. Connell TG, Rele M, Cowley D, Buttery JP, Curtis N. How reliable is a negative blood culture result? Volume of blood submitted for culture in routine practice in a children’s hospital. Pediatrics 2007;119(5):891–6. (In eng). DOI: 10.1542/peds.2006-0440.

9. Obiero CW, Gumbi W, Mwakio S, et al. Detection of pathogens associated with early-onset neonatal sepsis in cord blood at birth using quantitative PCR. Wellcome Open Res 2022;7:3. (In eng). DOI: 10.12688/wellcomeopenres.17386.2.

10. Velaphi SC, Westercamp M, Moleleki M, et al. Surveillance for incidence and etiology of early-onset neonatal sepsis in Soweto, South Africa. PLoS One 2019;14(4):e0214077. (In eng). DOI: 10.1371/journal.pone.0214077.

11. Fuchs A, Bielicki J, Mathur S, Sharland M, Van Den Anker JN. Reviewing the WHO guidelines for antibiotic use for sepsis in neonates and children. Paediatr Int Child Health 2018;38(sup1):S3–S15. (In eng). DOI: 10.1080/20469047.2017.1408738.

12. Labi AK, Enweronu-Laryea CC, Nartey ET, et al. Bloodstream Infections at Two Neonatal Intensive Care Units in Ghana: Multidrug Resistant Enterobacterales Undermine the Usefulness of Standard Antibiotic Regimes. Pediatr Infect Dis J 2021;40(12):1115–1121. (In eng). DOI: 10.1097/INF.0000000000003284.

13. Nebbioso A, Ogundipe OF, Repetto EC, et al. When first line treatment of neonatal infection is not enough: blood culture and resistance patterns in neonates requiring second line antibiotic therapy in Bangui, Central African Republic. BMC Pediatr 2021;21(1):570. (In eng). DOI: 10.1186/s12887-021-02911-w.

14. Zamarano H, Musinguzi B, Kabajulizi I, et al. Bacteriological profile, antibiotic susceptibility and factors associated with neonatal Septicaemia at Kilembe mines hospital, Kasese District Western Uganda. BMC Microbiol 2021;21(1):303. (In eng). DOI: 10.1186/s12866-021-02367-z.

15. Paulson JN, Williams BL, Hehnly C, et al. infection with frequent viral coinfection contributes to postinfectious hydrocephalus in Ugandan infants. Sci Transl Med 2020;12(563) (In eng). DOI: 10.1126/scitranslmed.aba0565.

16. Isaacs AM, Morton SU, Movassagh M, et al. Immune activation during. iScience 2021;24(4):102351. (In eng). DOI: 10.1016/j.isci.2021.102351.

17. Morton SU, Hehnly C, Burgoine K, et al. Paenibacillus Infection Causes Neonatal Sepsis and Subsequent Postinfectious Hydrocephalus in Ugandan Infants.. SSRN; 2022.

18. Drotar D, Olness K, Wiznitzer M, et al. Neurodevelopmental outcomes of Ugandan infants with human immunodeficiency virus type 1 infection. Pediatrics 1997;100(1):E5. (In eng). DOI: 10.1542/peds.100.1.e5.

19. Luna VA, King DS, Gulledge J, Cannons AC, Amuso PT, Cattani J. Susceptibility of Bacillus anthracis, Bacillus cereus, Bacillus mycoides, Bacillus pseudomycoides and Bacillus thuringiensis to 24 antimicrobials using Sensititre automated microbroth dilution and Etest agar gradient diffusion methods. J Antimicrob Chemother 2007;60(3):555–67. (In eng). DOI: 10.1093/jac/dkm213.

20. Ouyang J, Pei Z, Lutwick L, et al. Case report: Paenibacillus thiaminolyticus: a new cause of human infection, inducing bacteremia in a patient on hemodialysis. Ann Clin Lab Sci 2008;38(4):393–400. (In eng).

21. Di Micco R, Schneider M, Nüesch R. Postoperative Paenibacillus thiaminolyticus Wound Infection, Switzerland. Emerg Infect Dis 2021;27(7):1984–1986. (In eng). DOI: 10.3201/eid2707.203348.

22. Hunt B, Rogers C, Blais RM, Adachi K, Sathyavagiswaran L. Paenibacillus Sepsis and Meningitis in a Premature Infant: A Case Report. Am J Forensic Med Pathol 2021;42(1):96–98. (In eng). DOI: 10.1097/PAF.0000000000000610.

23. Movassagh M, Bebell LM, Burgoine K, et al. Vaginal microbiome topic modeling of laboring Ugandan women with and without fever. NPJ Biofilms Microbiomes 2021;7(1):75. (In eng). DOI: 10.1038/s41522-021-00244-1.

24. Richter CA, Wright-Osment MK, Zajicek JL, Honeyfield DC, Tillitt DE. Quantitative polymerase chain reaction (PCR) assays for a bacterial thiaminase I gene and the thiaminase-producing bacterium Paenibacillus thiaminolyticus. J Aquat Anim Health 2009;21(4):229–38. (In eng). DOI: 10.1577/H07-054.1.

25. Dhawan S, Singh R, Kaur R, Kaur J. A β-mannanase from Paenibacillus sp.: Optimization of production and its possible prebiotic potential. Biotechnol Appl Biochem 2016;63(5):669–678. (In eng). DOI: 10.1002/bab.1419.

26. Dsouza M, Taylor MW, Turner SJ, Aislabie J. Genome-based comparative analyses of Antarctic and temperate species of Paenibacillus. PLoS One 2014;9(10):e108009. (In eng). DOI: 10.1371/journal.pone.0108009.

27. Chauhan NS, Joseph N, Shaligram S, et al. Paenibacillus oleatilyticus sp. nov., isolated from soil. Arch Microbiol 2022;204(8):516. (In eng). DOI: 10.1007/s00203-022-03116-0.

28. da Silva FKL, de Sa Alexandre AR, Casas AA, et al. Increased production of chitinase by a Paenibacillus illinoisensis isolated from Brazilian coastal soil when immobilized in alginate beads. Folia Microbiol (Praha) 2022 (In eng). DOI: 10.1007/s12223-022-00992-3.

29. Kumar S, Ujor VC. Complete Genome Sequence of Paenibacillus polymyxa DSM 365, a Soil Bacterium of Agricultural and Industrial Importance. Microbiol Resour Announc 2022;11(6):e0032922. (In eng). DOI: 10.1128/mra.00329-22.

30. Lee M, Ten LN, Baek SH, Im WT, Aslam Z, Lee ST. Paenibacillus ginsengisoli sp. nov., a novel bacterium isolated from soil of a ginseng field in Pocheon Province, South Korea. Antonie Van Leeuwenhoek 2007;91(2):127–35. (In eng). DOI: 10.1007/s10482-006-9102-x.

31. Dornelles LV, Procianoy RS, Roesch LFW, et al. Meconium microbiota predicts clinical early-onset neonatal sepsis in preterm neonates. J Matern Fetal Neonatal Med 2022;35(10):1935–1943. (In eng). DOI: 10.1080/14767058.2020.1774870.

32. Dang T, Loll B, Müller S, et al. Molecular basis of antibiotic self-resistance in a bee larvae pathogen. Nat Commun 2022;13(1):2349. (In eng). DOI: 10.1038/s41467-022-29829-w.

33. DeLeon SD, Welliver RC. Paenibacillus alvei Sepsis in a Neonate. Pediatr Infect Dis J 2016;35(3):358. (In eng). DOI: 10.1097/INF.0000000000001003.

34. Zhang E, Lu H, Liu Q, et al. Paenibacillus assamensis in Joint Fluid of Man with Suspected Tularemia, China. Emerg Infect Dis 2018;24(8):1589–1591. (In eng). DOI: 10.3201/eid2408.180260.

35. Wenzler E, Kamboj K, Balada-Llasat JM. Severe Sepsis Secondary to Persistent Lysinibacillus sphaericus, Lysinibacillus fusiformis and Paenibacillus amylolyticus Bacteremia. Int J Infect Dis 2015;35:93–5. (In eng). DOI: 10.1016/j.ijid.2015.04.016.

36. Isaacman DJ, Karasic RB, Reynolds EA, Kost SI. Effect of number of blood cultures and volume of blood on detection of bacteremia in children. J Pediatr 1996;128(2):190–5. (In eng). DOI: 10.1016/s0022-3476(96)70388-8.

37. Kellogg JA, Ferrentino FL, Goodstein MH, Liss J, Shapiro SL, Bankert DA. Frequency of low level bacteremia in infants from birth to two months of age. Pediatr Infect Dis J 1997;16(4):381–5. (In eng). DOI: 10.1097/00006454-199704000-00009.

38. Hehnly C, Zhang L, Paulson JN, et al. Complete Genome Sequences of the Human Pathogen Paenibacillus thiaminolyticus Mbale and Type Strain P. thiaminolyticus NRRL B-4156. Microbiol Resour Announc 2020;9(15) (In eng). DOI: 10.1128/MRA.00181-20.

39. Messbarger N, Neemann K. Role of Anaerobic Blood Cultures in Neonatal Bacteremia. J Pediatric Infect Dis Soc 2018;7(3):e65–e69. (In eng). DOI: 10.1093/jpids/pix088.

40. Birgisson H, Steingrimsson O, Gudnason T. Kingella kingae infections in paediatric patients: 5 cases of septic arthritis, osteomyelitis and bacteraemia. Scand J Infect Dis 1997;29(5):495–8. (In eng). DOI: 10.3109/00365549709011861.

41. Yagupsky P, Dagan R, Howard CW, Einhorn M, Kassis I, Simu A. High prevalence of Kingella kingae in joint fluid from children with septic arthritis revealed by the BACTEC blood culture system. J Clin Microbiol 1992;30(5):1278–81. (In eng). DOI: 10.1128/jcm.30.5.1278-1281.1992.

42. Che G, Liu F, Chang L, Lai S, Teng J, Yang Q. Mycoplasma hominis Meningitis Diagnosed by Metagenomic Next-Generation Sequencing in a Preterm Newborn: a Case Report and Literature Review. Lab Med 2022 (In eng). DOI: 10.1093/labmed/lmac078.

43. Qin L, Li YH, Cao XJ, et al. Clinical metagenomic sequencing for rapid diagnosis of neonatal meningitis caused by Ureaplasma parvum: A case report. Medicine (Baltimore) 2022;101(4):e28662. (In eng). DOI: 10.1097/MD.0000000000028662.

44. Decroix V, Goudjil S, Kongolo G, Mammeri H. ‘Leptotrichia amnionii’, a newly reported cause of early onset neonatal meningitis. J Med Microbiol 2013;62(Pt 5):785–788. (In eng). DOI: 10.1099/jmm.0.051870-0.

45. Vitorino P, Varo R, Castillo P, et al. Sneathia amnii and Maternal Chorioamnionitis and Stillbirth, Mozambique. Emerg Infect Dis 2019;25(8):1614–1616. (In eng). DOI: 10.3201/eid2508.190526.

46. Kralik SF, Kukreja MK, Paldino MJ, Desai NK, Vallejo JG. Comparison of CSF and MRI Findings among Neonates and Infants with. AJNR Am J Neuroradiol 2019;40(8):1413–1417. (In eng). DOI: 10.3174/ajnr.A6134.

47. Xu M, Hu L, Huang H, et al. Etiology and Clinical Features of Full-Term Neonatal Bacterial Meningitis: A Multicenter Retrospective Cohort Study. Front Pediatr 2019;7:31. (In eng). DOI: 10.3389/fped.2019.00031.

48. Cromwell EA, Dube Q, Cole SR, et al. Validity of US norms for the Bayley Scales of Infant Development-III in Malawian children. Eur J Paediatr Neurol 2014;18(2):223–30. (In eng). DOI: 10.1016/j.ejpn.2013.11.011.

49. Pendergast LL, Schaefer BA, Murray-Kolb LE, et al. Assessing development across cultures: Invariance of the Bayley-III Scales Across Seven International MAL-ED sites. Sch Psychol Q 2018;33(4):604–614. (In eng). DOI: 10.1037/spq0000264.

50. Downie L, Armiento R, Subhi R, Kelly J, Clifford V, Duke T. Community-acquired neonatal and infant sepsis in developing countries: efficacy of WHO’s currently recommended antibiotics--systematic review and meta-analysis. Arch Dis Child 2013;98(2):146–54. (In eng). DOI: 10.1136/archdischild-2012-302033.

51. thaver D, Ali SA, Zaidi AK. Antimicrobial resistance among neonatal pathogens in developing countries. Pediatr Infect Dis J 2009;28(1 Suppl):S19–21. (In eng). DOI: 10.1097/INF.0b013e3181958780.

52. Dharmapalan D, Shet A, Yewale V, Sharland M. High Reported Rates of Antimicrobial Resistance in Indian Neonatal and Pediatric Blood Stream Infections. J Pediatric Infect Dis Soc 2017;6(3):e62–e68. (In eng). DOI: 10.1093/jpids/piw092.

53. C H, A S, P S, et al. Type IV pili is a critical virulence factor in clinical isolates of Paenibacillus thiaminolyticus. bioRxiv; 2022.

54. Sáez-Nieto JA, Medina-Pascual MJ, Carrasco G, et al. spp. isolated from human and environmental samples in Spain: detection of 11 new species. New Microbes New Infect 2017;19:19–27. (In eng). DOI: 10.1016/j.nmni.2017.05.006.

55. Chu SM, Hsu JF, Lai MY, et al. Risk Factors of Initial Inappropriate Antibiotic Therapy and the Impacts on Outcomes of Neonates with Gram-Negative Bacteremia. Antibiotics (Basel) 2020;9(4) (In eng). DOI: 10.3390/antibiotics9040203.

56. Apisarnthanarak A, Holzmann-Pazgal G, Hamvas A, Olsen MA, Fraser VJ. Antimicrobial use and the influence of inadequate empiric antimicrobial therapy on the outcomes of nosocomial bloodstream infections in a neonatal intensive care unit. Infect Control Hosp Epidemiol 2004;25(9):735–41. (In eng). DOI: 10.1086/502469.

57. thaden JT, Ericson JE, Cross H, et al. Survival Benefit of Empirical Therapy for Staphylococcus aureus Bloodstream Infections in Infants. Pediatr Infect Dis J 2015 (In eng). DOI: 10.1097/INF.0000000000000850.

58. Hsu JF, Chu SM, Wang HC, et al. Multidrug-Resistant Healthcare-Associated Infections in Neonates with Severe Respiratory Failure and the Impacts of Inappropriate Initial Antibiotic Therap. Antibiotics (Basel) 2021;10(4) (In eng). DOI: 10.3390/antibiotics10040459.

59. Kiwanuka J, Bazira J, Mwanga J, et al. The microbial spectrum of neonatal sepsis in Uganda: recovery of culturable bacteria in mother-infant pairs. PLoS One 2013;8(8):e72775. (In eng). DOI: 10.1371/journal.pone.0072775.

60. Popescu CR, Cavanagh MMM, Tembo B, et al. Neonatal sepsis in low-income countries: epidemiology, diagnosis and prevention. Expert Rev Anti Infect Ther 2020;18(5):443–452. (In eng). DOI: 10.1080/14787210.2020.1732818.

61. Sullins AK, Abdel-Rahman SM. Pharmacokinetics of antibacterial agents in the CSF of children and adolescents. Paediatr Drugs 2013;15(2):93–117. (In eng). DOI: 10.1007/s40272-013-0017-5.

62. Guardabassi L, Christensen H, Hasman H, Dalsgaard A. Members of the genera Paenibacillus and Rhodococcus harbor genes homologous to enterococcal glycopeptide resistance genes vanA and vanB. Antimicrob Agents Chemother 2004;48(12):4915–8. (In eng). DOI: 10.1128/AAC.48.12.4915-4918.2004.

63. tsai AL, Hsieh YC, Chen CJ, et al. Investigation of a cluster of Bacillus cereus bacteremia in neonatal care units. J Microbiol Immunol Infect 2022;55(3):494–502. (In eng). DOI: 10.1016/j.jmii.2021.07.008.

64. Ap GR, Daga MK, Mawari G, et al. Effect of Supplementation of Vitamin C and Thiamine on the Outcome in Sepsis: South East Asian Region. J Assoc Physicians India 2022;70(3):11–12. (In eng).

65. Na W, Shen H, Li Y, Qu D. Hydrocortisone, ascorbic acid, and thiamine (HAT) for sepsis and septic shock: a meta-analysis with sequential trial analysis. J Intensive Care 2021;9(1):75. (In eng). DOI: 10.1186/s40560-021-00589-x.

66. Sevransky JE, Rothman RE, Hager DN, et al. Effect of Vitamin C, Thiamine, and Hydrocortisone on Ventilator- and Vasopressor-Free Days in Patients With Sepsis: The VICTAS Randomized Clinical Trial. JAMA 2021;325(8):742–750. (In eng). DOI: 10.1001/jama.2020.24505.

67. Zhang L, Li S, Lu X, et al. Thiamine May Be Beneficial for Patients With Ventilator-Associated Pneumonia in the Intensive Care Unit: A Retrospective Study Based on the MIMIC-IV Database. Front Pharmacol 2022;13:898566. (In eng). DOI: 10.3389/fphar.2022.898566.

68. Li T, Bai J, Du Y, et al. Thiamine pretreatment improves endotoxemia-related liver injury and cholestatic complications by regulating galactose metabolism and inhibiting macrophage activation. Int Immunopharmacol 2022;108:108892. (In eng). DOI: 10.1016/j.intimp.2022.108892.

69. Raj KM, Baranwal AK, Attri SV, et al. Thiamine Status in Children with Septic Shock from a Developing Country: A Prospective Case-Control Study. J Trop Pediatr 2021;67(1) (In eng). DOI: 10.1093/tropej/fmaa107.

70. Donnino MW, Andersen LW, Chase M, et al. Randomized, Double-Blind, Placebo-Controlled Trial of Thiamine as a Metabolic Resuscitator in Septic Shock: A Pilot Study. Crit Care Med 2016;44(2):360–7. (In eng). DOI: 10.1097/CCM.0000000000001572.

71. Measelle JR, Baldwin DA, Gallant J, et al. Thiamine supplementation holds neurocognitive benefits for breastfed infants during the first year of life. Ann N Y Acad Sci 2021;1498(1):116–132. (In eng). DOI: 10.1111/nyas.14610.

72. Sambon M, Wins P, Bettendorff L. Neuroprotective Effects of Thiamine and Precursors with Higher Bioavailability: Focus on Benfotiamine and Dibenzoylthiamine. Int J Mol Sci 2021;22(11) (In eng). DOI: 10.3390/ijms22115418.

